# fNIRS-derived Neurocognitive Ratio as a Biomarker for Neuropsychiatric Diseases

**DOI:** 10.1101/2021.05.10.21256934

**Authors:** Ata Akın

## Abstract

**Significance:** Clinical use of fNIRS derived features has always suffered low sensitivity and specificity due to signal contamination from background systemic physiological fluctuations. This article provides an algorithm to extract cognition related features by eliminating the effect of background signal contamination; hence, improves the classification accuracy.

**Aim:** The aim in this study is to investigate the classification accuracy of an fNIRS derived biomarker based on global efficiency. To this end, fNIRS data were collected during a computerized Stroop Task from healthy controls, and patients with migraine, obsessive compulsive disorder, and schizophrenia.

**Approach:** Functional connectivity (FC) maps were computed from [HbO] time series data for Neutral, Congruent and Incongruent stimuli using the partial correlation approach. Reconstruction of FC matrices with optimal choice of principal components yielded two independent networks: Cognitive Mode Network (CM) and Default Mode Network(DM).

**Results:** Global Efficiency (*GE*) values computed for each FC matrix after applying principal component analysis yielded strong statistical significance leading to a higher specificity and accuracy. A new index, Neurocognitive Ratio (NCR), was computed by multiplying the Cognitive Quotients (*CQ*) and ratio of *GE* of CM to *GE* of DM. When mean values of *NCR* 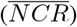 over all stimuli were computed, they showed high sensitivity (100%), specificity (95.5%), and accuracy (96.3%) for all subjects groups.

**Conclusions:** 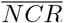 can reliable be used as a biomarker to improve the classification of healthy to neuropsychiatric patients.

## 1 Introduction

Although fNIRS has been around over 30 years now, its clinical efficacy and role are still being questioned due to its low specificity and sensitivity, especially in the area of neuropsychiatric diseases. Many researchers have been trying to improve its efficacy, sensitivity, and specificity in clinical settings by either improving its technology or the post processing analysis methods. Over the last 20 years, the richness of fNIRS data due to its ease and speed of data collection, non-invasiveness and access to local activity have become even more attractive to cognitive neuro-scientists in testing multitude of data processing and neuroscientific hypotheses. Strangely though, the brain does not work locally.^1–4^

So far, fNIRS researchers have focused more on the data analytics side than developing of novel technologies. We have enjoyed the availability of various fNIRS systems but at the cost of standardization of probe designs, data collection methodologies and even more on the data analytics.^5^ The lack of standardization on these issues has made it increasingly more difficult to compare the findings from different studies.^6^ Moreover, physics of photon migration through the layers of the head limits the specificity of the fNIRS device to cortical layers. Collected data become an amalgam of physiological activity from each layer the photon interacts with. Hence the data are known to be contaminated with background systemic physiological fluctuations that are undoubtedly correlated with the cognitive activity.^7,8^ The low specificity of the CW-fNIRS devices can be overcome with time resolved systems albeit at a greater cost and unease of data collection. Many novel data analysis methods have been proposed to extract the brain originated, task related data from the collected data.^9–11^ Still there is no consensus on how to approach the fNIRS data, leading to the unsettling yet quite accurate prediction of Drs. Quaresima and Ferrari:

> “*The prediction of the future directions of fNIRS for assessing brain function during human behavior in natural and social situations is not easy*.^5^”

So after spending more than 20 years on fNIRS research, I aimed to produce an analysis method (a pipeline of data analysis) that would avoid the pitfalls of the standardization issues. I recommend the following minimum hardware, data collection, and analysis requirements:

### 1. Hardware Requirements

1.1. Number of channels ≥ 8

1.2. Collect data from both hemispheres

1.3. Sampling rate ≥ 0.5 Hz per channel

1.4. Number of wavelengths ≥ 2

### 2. Task Requirements

2.1. Duration of collected data ≥ 10 minutes

2.2. Block stimulus with block duration ≥ 20 seconds

2.3. Stimulus type ≥ 2

2.4. Resting data ≈ 30 seconds

### 3. Analysis Requirements

3.1. HbO2 data

3.2. Avoid single channel data analysis

3.3. Prefer a dimensionless analysis (*i*.*e*. normalization of data)

3.4. Prefer a multichannel analysis (*i*.*e*. functional connectivity)

3.5. Prefer a metric that summarizes and captures the behaivour of multichannel data (*i*.*e*. functonal connectivity strength, global efficiency, *etc*.)

I have decided to consider these properties in proposing an improved post-processing approach to data obtained from fNIRS recordings over my last paper.^12^ The sole aim was to converge on a data analysis pipeline that will be accepted and adapted easily by *“fellow fNIRSians”*. The proposed algorithm should have a common denominator, a base, for all where anyone can build upon it. The algorithm aims to improve the statistical significance of the fNIRS findings; hence, the trust on the system. I aimed to boost the statistical significance of the Global Efficiency (*GE*) values; hence, the accuracy of classification of fNIRS findings in a set clinical data obtained in our group’s previous studies.

## 2 Methods and Materials

### 2.1 Subjects and Experimental Procedure

13 healthy subjects (6 female) at an average age of 26, 20 patients with migraine without aura (12 female) at an average age of 27, 26 patients with obsessive compulsive disorder (11 female) at an average age of 29, and 21 schizophrenia patients (10 female) at an average age of 28 participated in this study. The study protocol was approved by the Ethics Committee of Pamukkale University in 2008. Parts of these data were published by our group and coworkers.^13–22^ Consents were obtained from all subjects and they were all informed about the study before the experiment. Subjects were seated in a dimly illuminated insulated room and they were told to look at a computer screen placed in front of them.

Subjects responded to the computerized color word matching Stroop Task that involved 3 sets of stimuli: Neutral (N), Congruent (C), and Incongruent (I) stimuli. The task involved 15 N, 15 C and 15 I stimuli presented in blocks of five sequential stimuli. The inter stimulus interval was 4 seconds. The rest between each block was 20 seconds. The stimuli blocks were randomized for each subject. The subject was asked to respond with left or right mouse click depending on whether the stimulus was a match or not. The task started with a 30 seconds of rest and ended with a 30 seconds of rest.^21, 23^

### 2.2 fNIRS Equipment

The fNIRS system (NIROXCOPE 301) was developed at the Neuro-Optical Imaging Laboratory of Bogazici University (^14,24^). NIROXCOPE 301 has a sampling frequency of 1.77 Hz and it consists of a data acquisition unit, a data collecting computer and a flexible probe to place on the forehead of the subjects. The probe has a rectangular design housing 4 dual wavelength light emitting diodes (LED) emitting at 730 nm and 850 nm. Each LED is surrounded by 4 detectors placed 2.5 cm away from the center of the LEDs as seen in Figure **??**.

The validity of this probe design and its ability to detect brain tissue were discussed in our previous study^9^ as well as its efficacy in providing cognition related signals.^12,15 –17,22,25^

### 2.3 Analysis of the fNIRS Data

fNIRS data are known to be contaminated with systemic background fluctuations. Hence any correlation between two channels will be dominated by this common background signal. I have previously shown that using a high pass filter, computation of the inter-channel correlation via partial correlation provides a better insight into the underlying connectivity due to task. In my previous paper, I have outlined the analysis steps in detail.^12^ As a quick summary, I developed a signal processing pipeline to compute the functional connectivity matrices from [HbO] signals by using a partial correlation method, rather than the conventional pearson correlation analysis. Then these matrices were used to compute the global efficiency values. In this paper I propose an additional step between the FC matrices and GE computation by employing the principal component analysis. As a last step I propose two new biomarkers: Neurocognitive Index (*NCI*) and Neurocognitive Ratio (*NCR*). The details of the computation of these biomarkers are explained in Section 2.4 and the block diagram of the algorithm based is shown in Figure 1.

**Fig 1.**
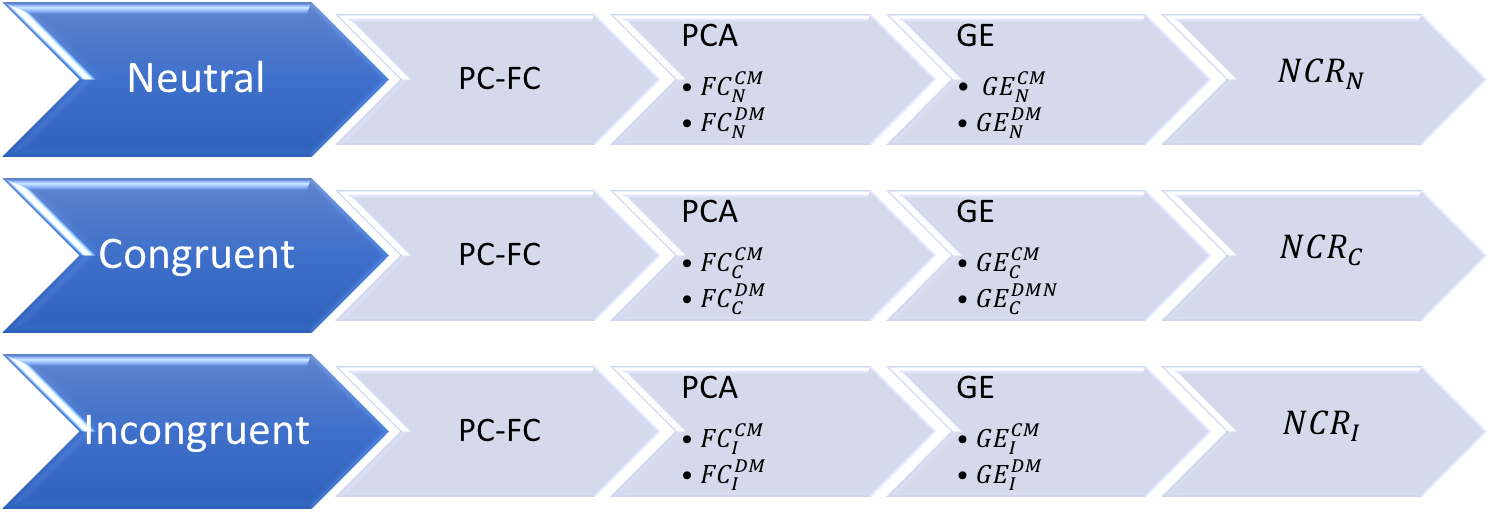
Block diagram of the *NCR* algorithm. CMN: Cognitive Mode Network, DMN: Default Mode Network

Statistical significance of the *GE* values were improved by adding principal component analysis (PCA) to the ℱ 𝒞 matrices.^26^ This paper will present the results of this PCA based FC analysis, hence called the FC-(PC)^2^: Functional connectivity analysis via PCA based PC.

#### 2.3.1 Functional Connectivity via PC

Partial correlation (PC) provides a relationship between two variables after removing the overlap from both variables. The partial correlation coefficient between any two channels (*i, j*) as *r*_*i,j*|*k*_ in the presence of a a common influencer (*k*) (*r*_*i,j*|*k*_) is as follows:^27^

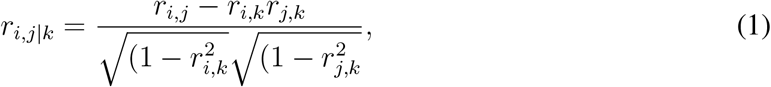

[HbO] data from each channel is passed through a high pass filter (butterworth, 8th order, *f*_*c*_ = 0.09 Hz) to obtain the 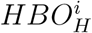. The regressor used in PC-based FC analysis is obtained by averaging this signal over all the channels. Hence 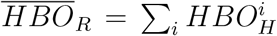 is used to regress out the systemic physiological affects from the correlation of the unprocessed [HbO] signals from two channels. Once the regressor 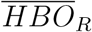 is computed, neutral (*N*), congruent (*C*), and incongruent (*I*) stimuli parts are segmented out individually and consolidated to form one individual time series from each of these stimuli. The functional connectivity matrices computed for individual time series are thus termed as ℱ𝒞_*N*_, ℱ𝒞_*C*_, ℱ𝒞_*I*_. This analysis is performed for each subject.

#### 2.3.2 Functional Connectivity via PCA based PC

Once the FC matrices are generated, I applied the PCA to the matrices. Since the matrices are 16 × 16, there are 16 principal components. An optimal choice for these components should improve the statistical significance of the *GE*_*i*_ values for computed from each ℱ𝒞_*i*_, (*i* being the stimulus type). The assumption in applying PCA to ℱ𝒞 matrices is the following:

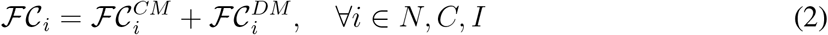

where 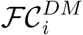 is the FC matrix of the Default Mode Network, while 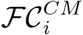 is the matrix for the Cognitive Mode Network. Since these two matrices *can be assumed to be linearly independent*, I used PCA to separate them. The choice of principal components (PCs) turned out to affect the statistical significance of the *GE* values computed after new ℱ𝒞 matrices were reconstructed from the chosen principal components. The expectation is the convergence to a subset of PCs that will yield the strongest significance for 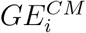 (as explained in Section 2.3.3)while no significance for the 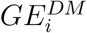.^28,29^ A set of PCs are used to reconstruct the new 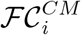. Once the best PC subset is found, this is determined as the CMN. Remaining PCs are then used to reconstruct the DMN matrices 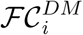, ∀*i* ∈ *N, C, I*. The expectation from these 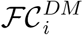 matrices is such that the t-statistics of the 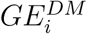, ∀*i* ∈ *N, C, I* will be low (no statistical significance). So the algorithm works as an optimization approach where the goal is to maximize the t-statistics (minimize the *p* value) of the 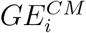, ∀*i* ∈ *N, C, I*.

#### 2.3.3 Global Efficiency

I decided to use the graph efficiency as a metric to quantify the information sharing efficiency of the 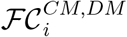 matrices. This approach is intrinsic to cognitive neuroscience where the aim is to investigate the neural correlates of cognition.^30–32^ Global efficiency is one of the many metrics of graph-based network analysis and has been used in brain connectivity studies.^12,33,–35^ We considered the channels as a set of vertices *V* and the partial correlation coefficients as assigned weights on the set of edges *E*, between vertices to construct an undirected complete weighted graph *G* = (*V, E*).^36–38^ ℱ𝒞 matrices are computed for each type o stimuli.

Global efficiency can be evaluated for a wide range of networks, including weighted graphs.^38^ Maximal possible global efficiency occurs when all edges are present in the network. The global efficiency value was computed by using the formulation of Latora and Marchiori’s,^39^ since it applies to work with weighted connectivity graphs. In this case, the global efficiency is:

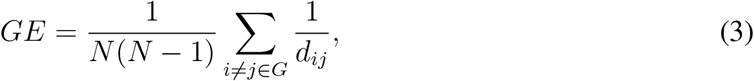

where *d*_*ij*_ is defined as the “smallest sum of the physical distances throughout all the possible paths in the graph from *i* to *j*”.^39^ For weighted graphs, stronger connection weights correspond to shorter lengths. Equation 3 generates values of *GE* in the range of [0, 1]. In this analysis, a threshold correlation value was determined iteratively for each individual FC matrix and the matrices were hard thresholded leaving only fewer non zero matrix entries. A review on the choice of such a threshold (*TH*) yielded a value of the highest 10 − 20% of all the entries. A sweep of the best *TH* that provides the highest statistical significance yielded specific *TH*_*s*_ values for different subject groups. Once the subject-optimized *TH* is found, the 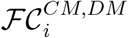 matrices are converted to binary matrices for computation of the 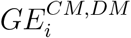 values by Eq’n 3 by the `efficiency.m` code from the Brain Connectivity Toolbox.^4^

### 2.4 Neurocognitive Index and Ratio

Cognitive quotient can be considered as a measure of level of cognitive effort exerted to fulfill a task. There are several indices, quotients and metrics proposed to assess this effort. Usually these are in the form of combinations of various neuropsychological task scores^40–42^ and sometimes in the form of physiological parameters.^43^ Cognitive load is a similar concept and many physiological measures have also been proposed to quantify this effort.^44^ The assumption for using physiological measures to quantify cognitive load is that brain, just like a muscle, has to produce some sort of a measurable physiological activity for a specific cognitive task.^45^ So the holly grail of neuroscience is to find this link between neurophysiological activity and psychological activity, also called the neurobiological basis of behavior.^46,47^ Hence, researchers have defined a new concept “neural efficiency” to quantify the level of efficiency of collaborative effort of the brain in solving a difficult cognitive task (For a complete review see^48^). Neural efficiency can be computed from physiological parameters like the heart rate variability, EEG measurements, fMRI recordings and recently from fNIRS findings.^49–51^ Researchers preferred to find a relationship between the neuropsychological data and neurophysiological data mostly in terms of correlation coefficients or regression analysis. The equation obtained transforms one finding to another one, consequently assumes a causal relationship.

I have decided to impose the duality principle in brain’s operations; hence, combined the behavioral findings with physiological findings in assessing the neurocognitive effort. I call this new combined metric the Neurocognitive Index (*NCI*). The analysis left at Section 2.3.3 continues with the calculation of the Neurocognitive Index (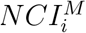 where *M* = *CM* or *DM*) with respect to stimulus as follows:

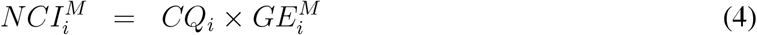

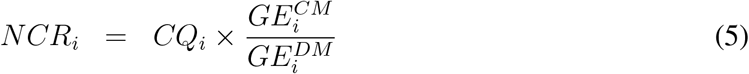

where *i* is the stimulus type. A further refinement of *NCI* is the Neurocognitive Ratio (*NCR*_*i*_). *NCR*_*i*_, as calculated from Equation 5, can be assumed to be a biomarker specific for each subject group (*i*.*e*. healthy controls, patients with migraine, OCD or Schizophrenia disorder). The underlying assumption in proposing this index as a biomarker is that, the global efficiency of a default mode network should be different than the global efficiency computed during a cognitive task and that the ratio of the two is an indicator of attention and inhibition control. In fact one can even hypothesize that an increased demand for inhibitory control can be associated with restructuring of the global network into a configuration that must be more optimized for specialized processing (functional segregation), more efficient at communicating the output of such processing across the network (functional integration), and more resilient to potential interruption (resilience). Thus, investigation of graph theoretical metrics under varying levels of inhibitory control can guide clinicians in classfying the severity of the disease as well as in prognosis.^46,52,53^

## 3 Results

### 3.1 Behavioral Results

The reaction times and accuracy rates for the subjects for all stimuli types are given in Figures 2(a) and 2(b). Reaction times are calculated by averaging the response times to all the responses, not just the correct answers.

**Fig 2.**
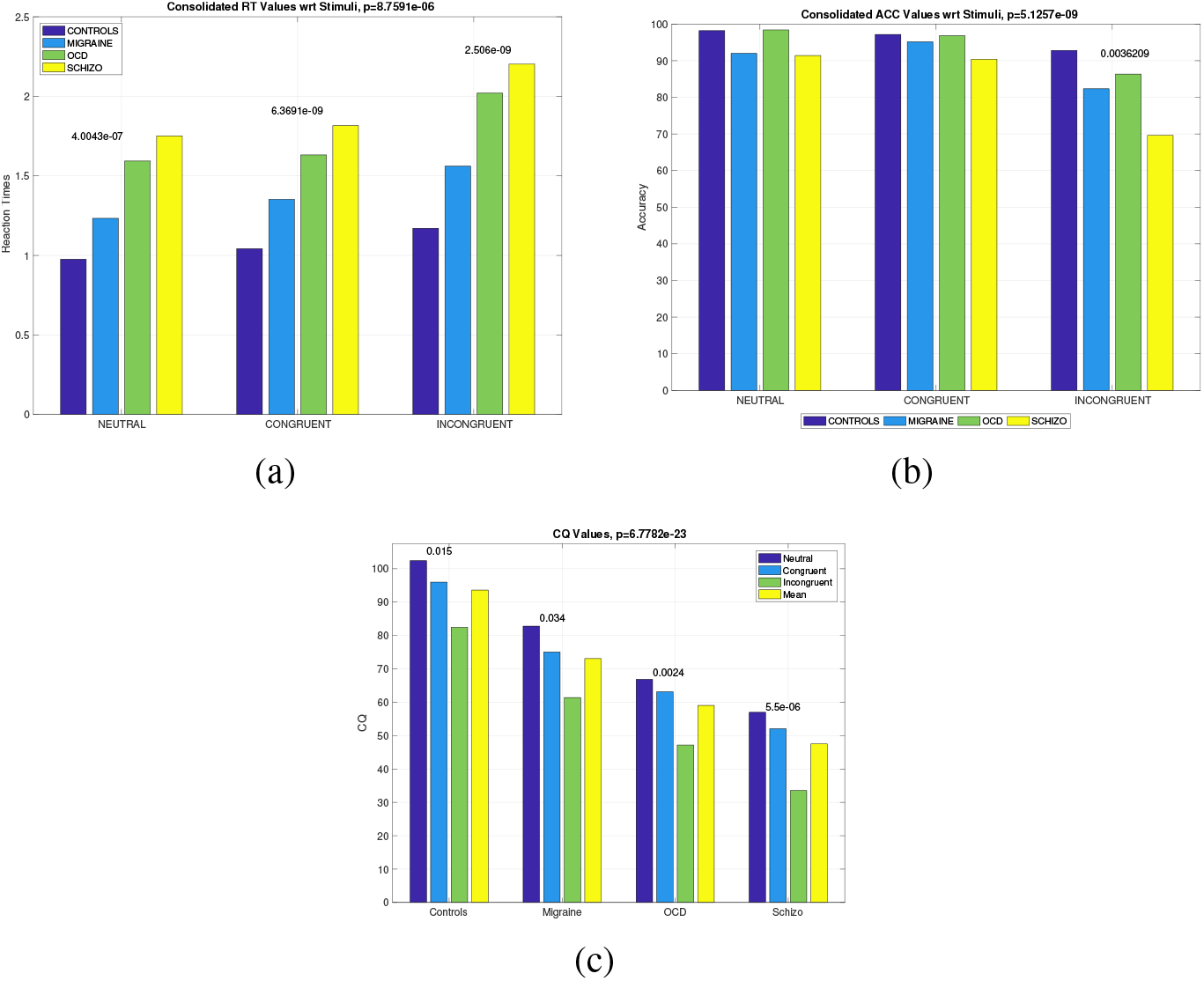
(a) *RT* values at the columns of Table 2 for all subjects across stimuli. *p* values are presented on top of bars. (b) *ACC* values at the columns of Table 3 for all subjects across stimuli. *p* values are presented on top of bars, (c) *CQ* values computed by Eq’n 6.

Two-way ANOVA for *RT* values yielded significant values for subject comparison (*p* = 0), and stimulus type (*p* = 0) and no interaction for *SUB*STIM* (*p* = 0.749). Accuracy is calculated by taking the ratio of total number correct answers to total number of questions. Two-way ANOVA for the *ACC* yielded significant values for subject comparison (*p* = 0), and stimulus type (*p* = 0) and no interaction for *SUB * STIM* (*p* = 0.1588).

A useful score for the behavioral results is the cognitive quotient (*CQ*) which is calculated by:

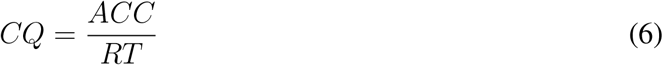

where *ACC* is the accuracy in percentage and *RT* is the reaction (response) time in seconds. *CQ* with respect to subjects and stimuli can be seen in Figure 2(c). Two-way ANOVA for *CQ* yielded significant values for subject comparison (*p* = 0), and stimulus type (*p* = 0) and no interaction for *SUB * STIM* (*p* = 0.9958). *CQ* can also be considered as a metric of cognitive load. In several studies such scores from different tests are linearly combined (sometimes with weights) to provide a stronger metric.

### 3.2 fNIRS Data Analysis

#### 3.2.1 GE Analysis

The computation of *GE*^*CM,DM*^ from the ℱ𝒞^*CM,DM*^ matrices yielded quite interesting dynamics as shown in Figures 3(a) and 3(b). Firstly, optimized *GE*^*CM*^ values showed a strong statistically significant difference within subjects, as displayed on top of the bars of Figure 3(a).

**Fig 3.**
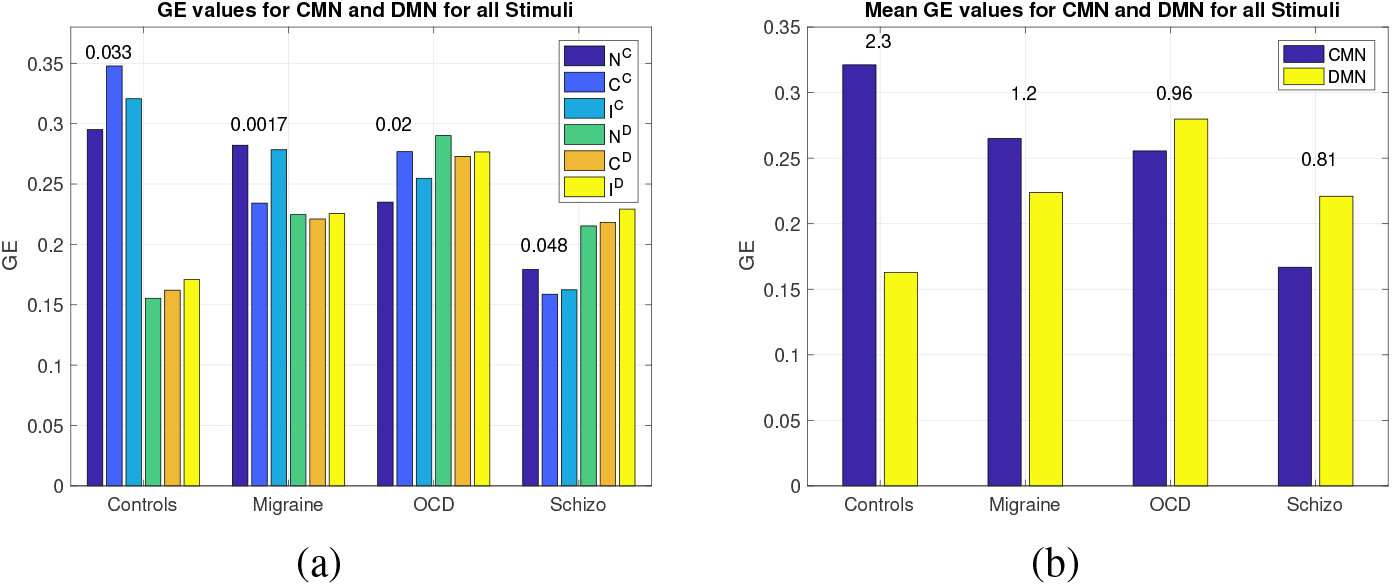
(a) *GE*^*CM,DM*^ values for all subjects across stimuli. *p* values are presented on top of bars, (b) Mean of *GE* values for all stimuli with respect to CMN and DMN. The numbers above the bars are the ratio of *GE* values for CMN to DMN

Secondly, both the *GE*^*CM*^ and *GE*^*DM*^ values are different between subject groups (as more evident from the means graph in Figure 3(b)). As the *GE*^*CM*^ value decreases from healthy controls to schizophrenics, the *GE*^*DM*^ increases. Higher value of *GE*^*CM*^ in controls over patients could mean that a healthy brain recruits a wider brain circuitry with more efficiency during a cognitive task, while diseased brain cannot. In contrast, higher values of *GE*^*DM*^ for diseased population might be due to a domination of the DMN leading to a lesser space for CMN; hence, the poor performance on cognition related activities. Figure 3(b) shows the ratio of mean of *GE*^*CM*^ */GE*^*DM*^ for each subject group. The ratio is in favor of healthy controls and significantly lower in diseased groups.

I should note that these values of *GE*^*CM,DM*^ depend heavily on the choice of optimal principal components and threshold values when computing the ℱ𝒞^*CM,DM*^ matrices.

#### 3.2.2 NCI and NCR Analysis

*NCI* and *NCR* values computed by Equations 4 and 5 elucidated strong statistical significance between healthy controls and rest of the diseased groups as seen in Figures 4(a) and 4(b).

**Fig 4.**
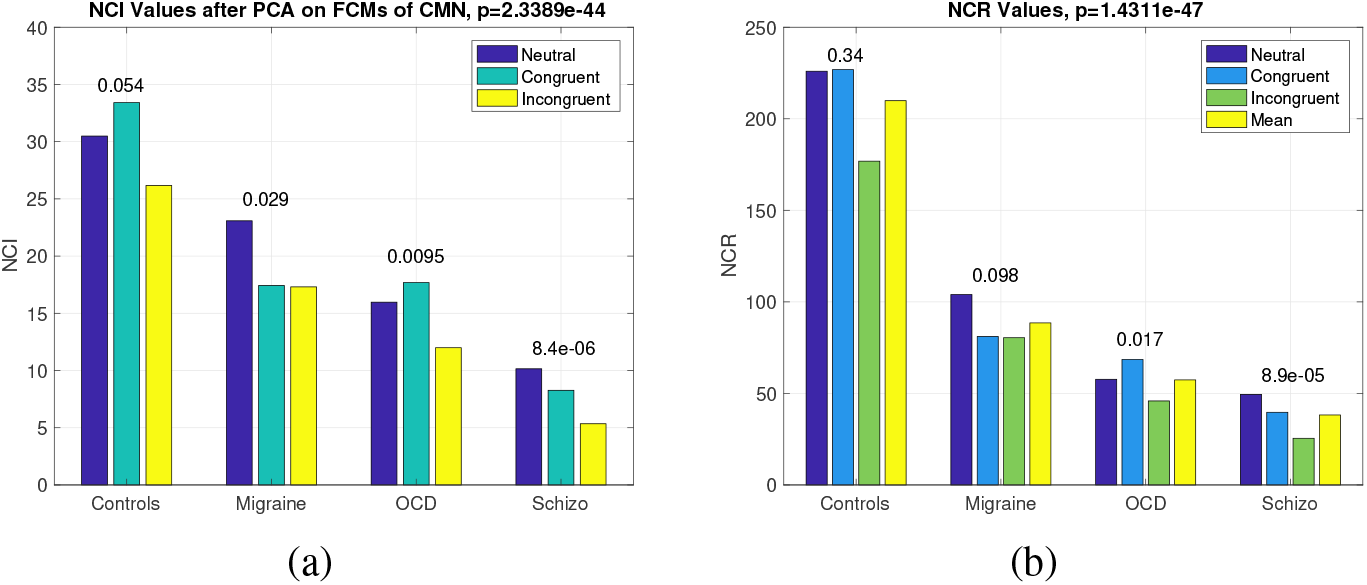
(a) *NCI* values as computed by Eq’n 4 for all stimuli across subjects, (b) *NCR* values as computed by Eq’n 5 for all subjects across stimuli. *p* values are presented on top of bars.

Two-way ANOVA for *NCI* yielded significant values for subject comparison (*p* = 0), and stimulus type (*p* = 0) and marginally significant interaction for *SUB * STIM* (*p* = 0.055). Two-way ANOVA for *NCR* yielded significant values for subject comparison (*p* = 0), and stimulus type (*p* = 0.001) and no interaction for *SUB * STIM* (*p* = 0.3575). As expected both the *NCI* and *NCR* values are highest for the healthy controls.

### 3.3 ROC Analysis

Receiver Operating Characteristics (ROC) provides a comparison of the accuracy of classification between the healthy controls and the rest of the cases. Figure 5(a) shows the results of the performance of classification among the parameters obtained in this paper. The whole aim to include fNIRS in such a study was to improve the classification accuracy of behavioral data from healthy and non-healthy persons.

**Fig 5.**
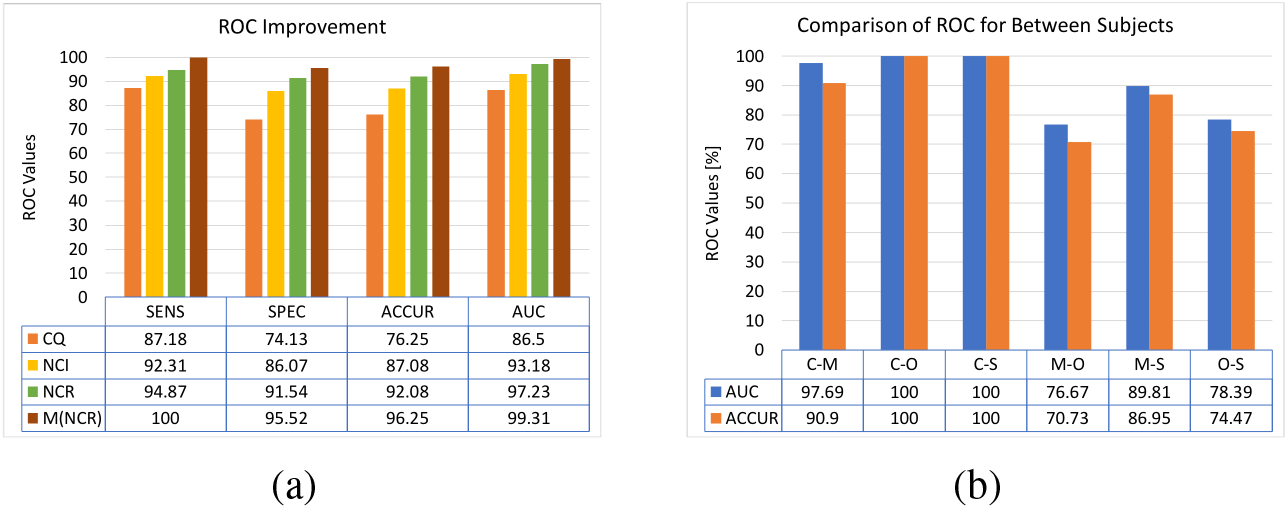
(a) ROC values computed for different features. Values are in percentages (%). SENS: Sensitivity, SPEC: Specificity, ACCUR: Accuracy, AUC: Area Under the Curve, 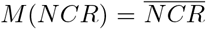, (b) ROC values between pairs of subjects, C: Controls, M: Migraineurs, O: OCD patients, S: Schizophrenia patients.

As promised the ROC values show a dramatic increase once the features derived from fNIRS findings are included alongside the behavioral findings. ROC values computed from the 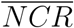 holds a great promise. The Sensitivity increases by 12.82%, Specificity by 21.39%, Accuracy by 20%, and AUC by 12.81% from the *CQ* based ROC values as displayed in Figure 5(a). One might wonder how the classification performance of 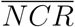 behaves between non-healthy subjects. That ROC analysis is given in Figure 5(b). *AUC* values for Controls vs. diseased patients are very high; hence, the sensitivity and specificity of the 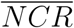 values are extraordinary in separating healthy from non-healthy brain.

## 4 Discussions

*fNIRSians* have long been search of a killer application that would secure the place of fNIRS in clinical settings. To this end, fNIRS have been applied to subjects of all ages and health conditions.^54–57^ Discussions regarding the limitations and ways to overcome these are a few yet they all have helped us redirect our efforts in proposing ever more innovative solutions. There are good reviews on the promise of fNIRS in neuropsychiatry.^5,47,54,56,58,–61^

This study is yet another one that proposes an algorithmic approach to the data analysis pipeline of fNIRS studies. The aim is to improve the clinical significance of the features extracted from fNIRS recordings so as to pledge an everlasting position of fNIRS in clinical settings. Only a handful studies investigated the differential diagnostic accuracy of fNIRS features.^62–66^ To start-off, I will address the specific expectations of any fNIRS based algorithmic approach for a clinical study:

**E1:** Provide clinically relevant information regarding brain physiology

**E2:** Provide strong specificity for clinical data

**E3:** Provide a better statistics than behavioral data alone

**E4:** Provide an easy and applicable/adoptable algorithm

fNIRS has been one of the few instruments that can provide insight to brain neurophysiology non-invasively and rapidly. Yet, these two offerings should match with the expectations listed above. Since fNIRS provides information regarding the cerebrovascular reactivity to cognitive or physiological stimuli, we expect that any measurement from patients with brain disorders should provide insight to neurobiology of the disease.

To address **E1**, fNIRS is famous for bestowing local hemodynamic activity. Moreover, *GE* extracted from the dynamic changes of such a local data elucidate the level of collaborative effort exerted during a cognitive task. So with number such as *GE* one can capture the hemodynamics of cognition.

To address **E2**, several groups reported medium to high accuracies for classification of fNIRS signals.^20,26,67,–70^ hese studies mostly included two groups: healthy controls and a diseased group. No multi-group comparison has been attempted with fNIRS, unlike fMRI.^26^ *NCR* derived from *CQ* and *GE* are shown to be highly specific for diseases.

To address **E3**, so far the statistical significance of behavioral data have been praised in many studies in classifying groups. fNIRS is expected to improve the statistics and this is what *NCR* offers. It gives a higher accuracy in separating healthy from diseased brain, much like a blood pressure monitor.

To address **E4**, I may be biased in thinking that the proposed approach is easy and applicable. It is more of an iterative approach than a theoretical one. Yet, the FC-(PC)^2^ seems to do the trick in separating the FC matrices.

### 4.1 On the Classification Accuracy of NCR

There have been several studies that investigated the classifier accuracy for schizophrenia patients. Yet, same cannot be said for migraine and OCD patients. Table 1 is a selection of such studies where search words: {*fNIRS, classification, schizophrenia, accuracy*} were used.

**Table 1.** Classification Performances of fNIRS studies

| Study | Diseased Group | Features | Classifier | Accuracy |
| --- | --- | --- | --- | --- |
| Chen et al.<sup>71</sup> | Schizophrenia | GLM based | SVM | 85% |
| Dadgostar et al.<sup>72</sup> | Schizophrenia | Time series | SVM | 87% |
| Einolou et al.<sup>20</sup> | Schizophrenia | Energy | SVM | 84% |
| Eken et al.<sup>67</sup> | SSD | FC based | SVM | 82% |
| Hahn et al.<sup>73</sup> | Schizophrenia | Time series | LOOCV | 76% |
| Ji et al.<sup>68</sup> | Schizophrenia | FC based | SVM | 89.7% |
| Shoushtarian et al.<sup>74</sup> | Chronic tinnitus | FC based | Naïve Bayes | 78.3% |
| Xu et al.<sup>69</sup> | ASD | Time series | CNN & LSTM | 93.3% |
| Yang et al.<sup>70</sup> | Schizophrenia | FC based | SVM | 84.67% |
| Yang et al.<sup>75</sup> | MCI | Time series | CNN | 98.61% |
| Yang et al.<sup>76</sup> | MCI | FC based | CNN | 95.81% |
| Yoo et al.<sup>77</sup> | MCI | Time series | SVM | 79.49% |

Most of the classification studies including schizophrenia patients reported a classification accuracy in the range of 76% to 89.7% as seen in Table 1. This study achieves at a 100% accuracy score for schizophrenia patiens as seen in Figure 5(b). The strength of this value is inherent to the computation of the 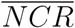 where behavioral and physiologic data are fused.

Interestingly there are not many studies of other psychiatric diseases. Still there are pioneering works on autism spectrum disorder,^61^ obsessive compulsive disorder,^59^ depression, and migraine.^14^ There is always those who have not lost faith in fNIRS.^47,60^ A very hopeful study by Ehlis et al.^58^ points us to the right direction:

> *“*…*future studies should also focus on the usefulness of fNIRS as a supportive tool for choosing the most promising treatment approach for a specific patient. Using fNIRS, neurophysiological markers that might predict treatment outcomes (and may thus be relevant for personalized medicine) could be easily identified*.*”*^*58*^

Several studies actually achieved this ambitious goal set by Ehlis et al. For a good review of use of fNIRS in psychiatry, please see,^58,78,79^specifically in autism,^61^ its role in neurofeedback,^80^ in pain,^81^ and in neurology.^82^ Only a handful of them are listed in Table 1. This study is the first in several aspects: 1) to show a high specificity of fNIRS for various types of neuropsychiatric diseases (more than 2), 2) in providing an fNIRS derived biomarker (namely the *NCR*) with very high accuracy that is also clinically relevant, and 3) in that it does not attempt to find a correlation between behavioral data and physiological data, rather it combines them since behavior cannot be produced without physiological activation.

## 5 Conclusion

> *“A science of the relations of mind and brain must show how the elementary ingredients of the former correspond to the elementary functions of the latter*.*”*^83^

This study is an extension of a previous work where I proposed a partial correlation based functional connectivity matrix computation. Separating the FC into a cognitive and default mode network led to ratio of the GE value calculated from these two matrices. This ratio was then multiplied with the cognitive quotient which is a direct measure of cognitive load. Therefore, I was able to generate a new biomarker, neurocognitive ratio (*NCR*). The statistical value of this biomarker was evaluated for its classification power. The results are all in favor of this biomarker. So we might conclude that *fNIRS derived NCR* is a strong candidate as a biomarker for neuropsychiatric diseases. It can safely be used in diagnosis and prognosis.

## Data Availability

Data and codes can be provided upon request to the author when found appropriate

## Acknowledgments

This project was sponsored by a grant from TUBITAK Project No: 108S101 and Bogazici University Research Fund (BURF Project No: 5106). Authors wish to thank Deniz Nevsehirli for his contributions to fNIRS optode development, Dr. Nermin Topaloĝu and Dr. Sinem Burcu Erdoĝan for data collection.

## Appendix

The following tables present the values of the figures in the manuscript.

### 6.1 Behavioural Data

**Table 2.** Reaction Times in milliseconds (mean ± standard deviation). Number of Subjects are given in parantheses

| Group | NS | CS | ICS | p-value |
| --- | --- | --- | --- | --- |
| Controls | 975.7 ± 134.7 | 1041.8 ± 190.8 | 1170.6 ± 227.6 | 0.0375* |
| Migraineurs | 1233.0 ± 394.6 | 1352.2 ± 345.9 | 1560.7 ± 576.8 | 0.0748 |
| OCD | 1593.1 ± 453.3 | 1631.5 ± 400.7 | 2019.7 ± 512.7 | 0.006* |
| Schizo | 1750.9 ± 458.1 | 1815.5 ± 323.4 | 2202.2 ± 346.4 | < 0.0001* |
| p-value | < 10<sup>-7</sup> | < 10<sup>-9</sup> | < 10<sup>-9</sup> | 0 |

**Table 3.** Accuracy rates in percentages (mean ± standard deviation).

| Group | NS | CS | ICS | p-value |
| --- | --- | --- | --- | --- |
| Controls (13) | 98.2 ± 2.2 | 97.2 ± 4.7 | 92.8 ± 7.4 | 0.0297* |
| Migraineurs | 92.0 ± 18.2 | 95.2 ± 8.4 | 82.3 ± 24.9 | 0.0815 |
| OCD | 98.4 ± 2.5 | 96.8 ± 4.4 | 86.3 ± 17.5 | < 0.001* |
| Schizo | 91.4 ± 13.9 | 90.4 ± 13.7 | 69.6 ± 21.4 | < 0.00001* |
| p-value | < 10<sup>-7</sup> | < 10<sup>-9</sup> | < 10<sup>-8</sup> | 0 |

Hence the following Table 4 is formed from Tables 2 and 3

**Table 4.** CQ values are given as (mean ± standard deviation).

| Group | NS | CS | ICS | p-value |
| --- | --- | --- | --- | --- |
| Controls | 102.42 ± 14.15 | 96.00 ± 16.87 | 82.44 ± 19.19 | 0.0147* |
| Migraineurs | 82.80 ± 27.50 | 75.04 ± 19.68 | 61.41 ± 28.62 | 0.0341* |
| OCD | 66.85 ± 19.49 | 63.12 ± 16.49 | 47.18 ± 19.44 | 0.0024* |
| Schizo | 56.99 ± 19.77 | 52.05 ± 14.62 | 33.53 ± 15.19 | < 0.00001* |
| p-value | < 10<sup>-7</sup> | < 10<sup>-9</sup> | < 10<sup>-8</sup> | 0 |

### 6.2 GE Values

**Table 5.** 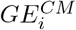 values computed from functional connectivity matrix of HbO data (mean ± standard deviation).

| Group | NS | CS | ICS | p-value |
| --- | --- | --- | --- | --- |
| Controls | $0.2951 \pm 0.0482$ | $0.3478 \pm 0.0543$ | $0.3207 \pm 0.0634$ | 0.0326 |
| Migraineurs | $0.2821 \pm 0.0275$ | $0.2342 \pm 0.0321$ | $0.2784 \pm 0.0287$ | 0.0017 |
| OCD | $0.2350 \pm 0.0662$ | $0.2768 \pm 0.0549$ | $0.2548 \pm 0.0549$ | 0.0197 |
| Schizo | $0.1793 \pm 0.0527$ | $0.1587 \pm 0.0701$ | $0.1624 \pm 0.0635$ | 0.0482 |
| p-value | $< 0.0001$ | $< 0.0001$ | $< 0.0001$ | 0 |

**Table 6.** 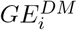 values computed from functional connectivity matrix of HbO data (mean ± standard deviation).

| Group | NS | CS | ICS | p-value |
| --- | --- | --- | --- | --- |
| Controls | $0.2951 \pm 0.0459$ | $0.3478 \pm 0.0463$ | $0.3207 \pm 0.0541$ | 0.032 |
| Migraineurs | $0.2515 \pm 0.0436$ | $0.2423 \pm 0.0481$ | $0.2466 \pm 0.0444$ | 0.8146 |
| OCD | $0.2204 \pm 0.0393$ | $0.2478 \pm 0.0471$ | $0.2502 \pm 0.0499$ | 0.0711 |
| Schizo | $0.2464 \pm 0.0513$ | $0.2232 \pm 0.0414$ | $0.2315 \pm 0.0593$ | 0.257 |
| p-value | 0.1065 | 0.1404 | 0.6088 | 0.4171 |

### 6.3 NCR Values

**Table 7.** *NCR* values computed from NE and GE (mean ± standard deviation).

| Group | NS | CS | ICS | p-value |
| --- | --- | --- | --- | --- |
| Controls | 225.97 ± 125.93 | 226.84 ± 80.88 | 176.85 ± 80.35 | 0.342 |
| Migraine | 104.00 ± 40.53 | 81.09 ± 28.04 | 80.41 ± 45.20 | 0.098 |
| OCD | 57.68 ± 23.92 | 68.54 ± 27.39 | 45.88 ± 22.78 | 0.017 |
| Schizo | 49.48 ± 25.05 | 39.68 ± 15.37 | 25.48 ± 14.43 | $< 10^{-5}$ |
| p-value | $< 10^{-10}$ | $< 10^{-10}$ | $< 10^{-10}$ | 0 |

**Table 8.**
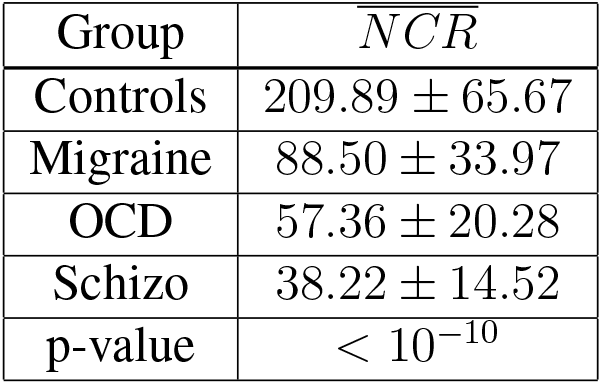
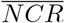 values computed from Table 7 (mean ± standard deviation).

| Group | $\overline{NCR}$ |
| --- | --- |
| Controls | 209.89 ± 65.67 |
| Migraine | 88.50 ± 33.97 |
| OCD | 57.36 ± 20.28 |
| Schizo | 38.22 ± 14.52 |
| p-value | $< 10^{-10}$ |

